# Safety and Efficacy of Adjunctive Intra-arterial Adenosine Following Successful Endovascular Thrombectomy in Patients with Large Vessel Occlusion Acute Ischemic Stroke (REACT) Trial: Rationale and Design

**DOI:** 10.64898/2026.01.06.26343497

**Authors:** Xianhua Hou, Yuxuan He, Xi Chen, Gaoming Li, Thanh N Nguyen, Jeffrey L. Saver, Duolao Wang, Wenjie Zi, Zhenhua Zhou, REACT investigators

**Affiliations:** Department of Neurology, The First Affiliated Hospital, Army Medical University, Chongqing, China; Department of Neurology, The Second Affiliated Hospital, Army Medical University, Chongqing, China; Departments of Radiology & Neurology, Boston Medical Center, Boston, MA, USA; Department of Neurology and Comprehensive Stroke Center, David Geffen School of Medicine, University of California, Los Angeles, Los Angeles, California; Global Health Trials Unit Liverpool School of Tropical Medicine Liverpool UK

**Keywords:** Clinical trial, Adenosine, Mechanical thrombectomy, Endovascular thrombectomy, Acute ischemic stroke, Large vessel occlusion

## Abstract

**Background:** Among patients with acute ischemic stroke (AIS) secondary to large vessel occlusion (LVO) who undergo successful reperfusion following endovascular thrombectomy (EVT), only one-third are disability-free at 90 days, which may be related to persistent microvascular hypoperfusion after thrombectomy known as the “no-reflow phenomenon”. Adenosine is administered to prevent percutaneous coronary intervention (PCI)-related no-reflow through microvasculature dilation and neutrophil-mediated inflammation modulation. However, its role in in the setting of AIS has not been clearly elucidated.

**Objective:** To evaluate the safety and efficacy of adjunctive intra-arterial adenosine following successful EVT in LVO patients.

**Methods and design:** In this multicenter, open-label, randomized, phase 2 trial, we evaluated the safety and efficacy of adjunctive intra-arterial adenosine following successful EVT in AIS patients. Up to 160 eligible stroke patients with anterior intracranial large vessel occlusion presenting within 24 hours from symptom onset (time last known well) are planned to be consecutively randomized. The primary outcome was the shift in the distribution of mRS scores at 90 days. Safety outcomes included symptomatic intracranial hemorrhage (sICH) within 48 hours and mortality at 90 days.

**Discussions:** This pivotal trial will provide first-hand data on the efficacy and safety of adjunctive intra-arterial adenosine following successful EVT in patients with acute ischemic stroke due to LVO.

**Trial registry number:** ChiCTR2400092051 (www.chictr.org.cn).

## Introduction

Stroke poses a significant global health burden as a leading cause of morbidity and mortality^1^. Despite endovascular thrombectomy (EVT) being the gold standard for acute ischemic stroke due to large vessel occlusion (LVO), many patients still suffer poor outcomes even after successful reperfusion^2^. The no-reflow phenomenon is a key contributor to this issue, with its pathophysiology involving multiple factors^3^. Firstly, distal emboli from proximal thrombus fragments and in - situ thrombus formation due to platelet, leukocyte, and fibrinogen stasis and deposition can hinder revascularization. Secondly, pericyte contraction, triggered by intracellular calcium overload, excitotoxicity, and oxidative/nitrosative stress during ischemia and even after successful revascularization, causes capillary narrowing and incomplete microcirculatory perfusion. Thirdly, ischemia - reperfusion induces leukocyte chemotaxis, aggregation, and activation, leading to excessive inflammatory responses that alter microvascular beds and hemorheology.

Although intra-arterial thrombolysis (IAT) has potential to improve functional outcomes by dissolving residual thrombi after endovascular recanalization^4–7^, recent RCTs (POST-TNK^5^, POST-UK^6^, ATTENTION-IA^7^) indicated that adjunctive IAT did not significantly increase the likelihood of freedom from disability. Furthermore, ATTENTION-IA indicated that intra-arterial tenecteplase increased symptomatic intracranial hemorrhage risk^7^. These findings indicate that post-thrombectomy IAT, by targeting residual micro-thrombi, is insufficient to resolve the no-reflow phenomenon; future research must explore additional mechanisms underlying persistent hypoperfusion. Recent experimental findings demonstrating the reversible nature of no-reflow offer promising prospects for a novel treatment paradigm in ischemic stroke^8^. Studies in animal models show that interventions targeting the cerebral vasculature—such as administering pericyte relaxants (e.g., adenosine)^9^ and the oxygen radical scavenger N-tert-butyl-α-phenylnitrone^10^, or depleting neutrophils via anti-Ly6G antibody^11^ to reduce capillary stalling—can effectively reinstitute capillary flow. These approaches not only prevent no-reflow but also lead to reduced infarct size and improved neurological outcomes.

Adenosine, a crucial endogenous purine nucleoside, has protective effects in the nervous system^12^. It works by activating purine receptors on vascular endothelial cells, inducing microarterial smooth muscle relaxation^13^. This leads to microvascular dilation, inhibition of inflammatory mediator expression and secretion, and platelet aggregation suppression. In brain ischemia, adenosine levels rise rapidly, offering neuroregulation and protection against excitotoxicity^14^. In the cardiovascular field, adenosine has been successfully used to treat no - reflow in acute coronary syndrome patients undergoing percutaneous coronary intervention. However, its application in acute anterior circulation large - vessel - occlusive stroke patients post - mechanical thrombectomy remains unexplored.

Drawing on cardiovascular experience, we hypothesize that administering adenosine after EVT in anterior circulation LVO stroke patients may improve microcirculation without increasing hemorrhagic transformation risks, leading to better prognosis. To test this hypothesis, we conduct an open-label, randomized, multicenter, exploratory trial to investigate the efficacy and safety of intra-arterial adenosine following successful thrombectomy in patients with acute ischemic stroke due to LVO within 24h of symptom onset.

## Methods

### Study design

The REACT trial is an investigator-initiated, randomized, open-label, blinded outcome assessment, phase 2 trial (Figure 1). This trial was registered at www.chictr.org.cn (ChiCTR2400092051). The trial is designed in compliance with the Declaration of Helsinki. The protocol was approved by the ethics committee of Chongqing Southwest Hospital, Army Medical University, and all participating centers. Data will be made available upon reasonable request to the corresponding author. This study includes 15 stroke centers in China. To qualify for participation in this trial, study centers will be mandated to have performed a minimum of 50 endovascular procedures each year. Additionally, all neurointerventionists involved will be required to have over five years of experience in cerebrovascular interventions, along with a track record of having performed at least 50 cases of mechanical thrombectomy using stent retriever devices annually. All endpoints of this study will be evaluated by independent investigators who are unaware of the treatment allocation and the actual treatment received. The trial flowchart is in Figure 2.

**Figure 1.**
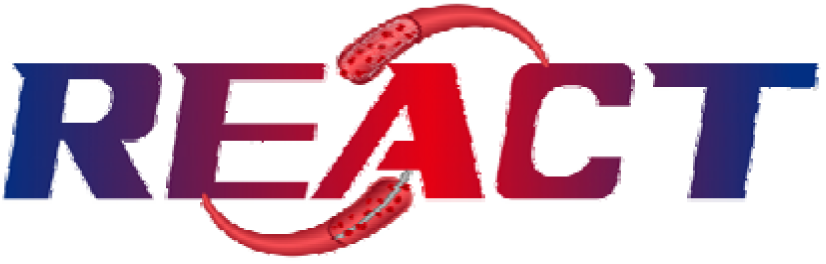
REACT trial logo.

**Figure 2.**
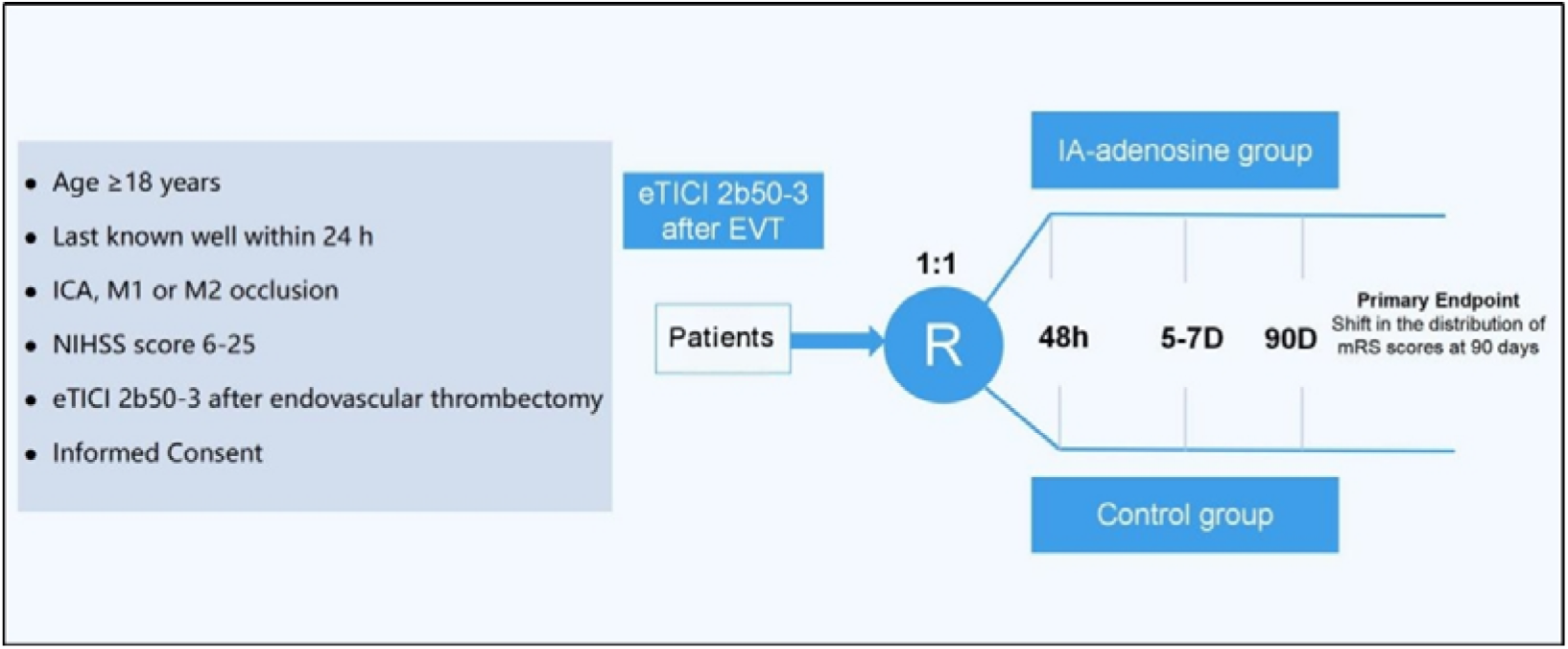
Flow chart of the REACT trial.

### Participant population

#### Inclusion criteria include

1. Age ≥18 years;
2. Presenting with acute ischemic stroke (AIS) with symptoms within 24 hours from time last known well;
3. Large vessel occlusive stroke in the anterior circulation confirmed by computed tomography angiography/magnetic resonance angiography, including intracranial segment of the internal carotid artery, middle cerebral artery M1 or M2 segment;
4. Baseline National Institutes of Health Stroke Scale (NIHSS) score: 6≤ NIHSS ≤ 25;
5. Successful reperfusion (eTICI 2b50-3);
6. Written informed consent signed by patient or their family member.

### Exclusion criteria

1. Intracranial hemorrhage confirmed by computed tomography or magnetic resonance imaging;
2. Prestroke mRS score ≥ 2;
3. Intraprocedural digital subtraction angiography (DSA) suggests vessel penetration, dissection, or extravasation of contrast medium;
4. Pregnant or lactating patients;
5. Allergic to contrast agent or Adenosine;
6. Degree II or III atrioventricular block or sick sinus syndrome (except for patients with artificial pacemakers);
7. Patients with known or estimated pulmonary disease with bronchial stenosis or bronchospasm (e.g., asthma);
8. Systolic pressure greater than 185 mmHg or diastolic pressure greater than 110 mmHg despite blood pressure lowering treatment;
9. Blood glucose < 2.8 mmol/L (50 mg/dl) or > 22.2 mmol/L (400 mg/dl);
10. Known bleeding tendency (including but not limited to): coagulation factor deficiency disease or platelets < 90*10^9/L or INR >3.0;
11. Patients on chronic hemodialysis and severe renal insufficiency (glomerular filtration rate < 30 ml/min or blood creatinine > 220μmol/L (2.5mg/dl));
12. Any terminal illness with a life expectancy of less than 6 months;
13. History of intracranial hemorrhage in the last 1 month;
14. Intracranial aneurysm, arteriovenous malformation;
15. Brain tumors with occupying effect on imaging;
16. Unlikely to be available for 90-day follow-up;
17. Current participation in another clinical trial;

### Randomization

Eligible patients were randomly assigned in a 1:1 ratio to the adenosine group or the control group via a real-time internet-based system (lcsjcj.xnyy.cn). Minimization randomization was stratified based on two parameters: patient age (<70 vs ≥70 years) and admission NIHSS score (<15 vs ≥15).

### Study Intervention

Both treatment groups will undergo EVT, which includes techniques such as stent retrievers, aspiration, balloon angioplasty, stenting, or any combination. After randomization, patients in the adenosine group will receive intra-arterial adenosine (120 ug as fast bolus followed by 2 mg given in 10 ml of saline over 2 min as slow bolus) through a distal access catheter or microcatheter located proximal to the original occlusion (eg, internal carotid artery, M1, or M2). Patients allocated to the control group will terminate the procedure without further intra-arterial adenosine.

All enrolled patients will be monitored in the acute stroke unit and can be admitted to the intensive care unit if necessary. All enrolled patients will undergo standardized medical treatment management and subsequent secondary preventive medication according to the Chinese Guidelines for Endovascular Treatment of Acute ischemic Stroke 2023^15^.

### Efficacy Endpoints

The primary end point is the distribution of the modified Rankin Scale(mRS) at 90 days after randomization.

### The secondary end points include

1. proportion of mRS score 0 to 1 at 90 (±7) days;
2. proportion of mRS score 0 to 2 at 90 (±7) days;
3. proportion of mRS score 0 to 3 at 90 (±7) days;
4. NIHSS change from baseline at 5-7 days or at discharge if earlier;
5. European Quality Five-Dimension scale score at 90 (±7) days.

### Safety Endpoints

The primary safety endpoints are mortality at 90 (±7) days, symptomatic intracranial hemorrhage (sICH) within 48h according to the modified Heidelberg Bleeding Classification^16^.

### Prespecified Exploratory Analyses

Additional prespecified exploratory analyses will assess the effect of the adenosine on plasma proteomics and markers of blood-brain barrier breakdown in 30 patients (15 in the adenosine group and 15 in the control group) from two centers (Dali People’s Hospital and Southwest Hospital). Plasma samples were collected 24 h after EVT and prepared by centrifuging EDTA-treated whole blood at 2,000×g for 10 min at 4℃. Plasma Proteomic analyses were performed using Olink Reveal assay. Markers of blood-brain barrier were evaluated using ELISA.

### Blinding and masking

Each participating site will allocate one or more physicians to perform follow-up evaluations at 24-hour intervals, between five to seven days post-treatment, or at the time of discharge if it occurs sooner, along with a 90-day assessment. It is imperative that these physicians are not involved in the subject’s initial treatment allocation to ensure their blinding to the treatment assignment.

### Data safety monitoring board

The independent Data and Safety Monitoring Committee (DSMB) will be set up, responsible for independently reviewing safety and efficacy data. The DSMB regularly reviews the study data added during the trial and advises the sponsor on the continued safety of the subjects and those not yet included in the study. The DSMB advises on the trial sponsor on the study’s scientific validity and overall conduct.

### Sample size

This study is a prospective, multicenter, open-label, randomized, controlled, end-point blind, exploratory phase 2 trial without formal sample size calculation. The planned initial sample size of this study is 160 patients, with 80 patients in the adenosine treatment group and 80 patients in the control group after EVT.

### Statistical analyses

Baseline demographic and clinical characteristics will be described using summary statistics. Continuous variables will be reported as medians with interquartile ranges (IQRs), while categorical variables will be expressed as counts and percentages. The primary endpoint will be assessed using the generalized odds ratio (genOR) with corresponding 95% CIs. The EQ-5D-5L and changes in NIHSS score will be using win ratio method. Binary outcomes and mortality will be compared between groups using a modified Poisson regression model with robust variance estimation and a Cox proportional hazards model, respectively. All analyses will be adjusted for age, NIHSS score, prestroke mRS score, ASPECTS, intravenous thrombolysis, occlusion site, and time to randomization. Both crude and adjusted effect estimates based on inverse probability of treatment weighting (IPTW) will be presented for each outcome. Subgroup analyses of the primary endpoint will be conducted across ten groups: age, sex, NIHSS score, prestroke mRS score, ASPECTS, time to randomization, intravenous thrombolysis, stroke etiology, occlusion location, and eTICI grade. Primary data analyses will be based on the intention-to-treat principle. The per-protocol analyses will also be performed as supplemental analyses. All statistical analyses will be described in detail in the statistical analysis plan which will be finalized before the study database lock, and will be performed using SAS version 9.4 (SAS Institute) and R version 4.4.0 or higher (R Foundation for Statistical Computing). The reporting of trial results will conform to the Consolidated Standards of Reporting Trials guidelines for randomized trials.

### Ethical considerations

The study adheres to the ethical guidelines outlined in the Declaration of Helsinki. The protocol was approved by the Ethics Committee of the First Affiliated Hospital of the Army Medical University and all participating centers. Any modifications to the trial protocol will only be executed after obtaining further approval from the ethics committee.

## Discussion

Currently, mechanical thrombectomy is the optimal treatment option for most acute large vessel occlusive ischemic stroke^17^. Despite advancements in thrombectomy devices and workflow processes, outcomes for patients with LVO remain suboptimal. The HERMES study indicated that one-third of the patients could not benefit from the treatment despite successful reperfusion^18^. The microcirculation reperfusion disorder may persist even if the large blood vessel recanalizes. In experimental animal models of cerebral ischemia/reperfusion, capillary occlusion, perivascular space obstruction, or distal microembolism from a proximal thrombus may prevent complete reperfusion of the distal microcirculation. The oxidized and nitrocompounds produced by arteriolar smooth muscle cells may also lead to microcirculation vasoconstriction and hypoperfusion^19^. These mechanisms also occur in the cerebral microvasculature of patients undergoing mechanical thrombectomy. Recently, Intra-arterial thrombolysis (IAT) adjunct to EVT has been applied to dissolve microthrombi and improve microcirculatory flow. Several RCTs trial^4–6^ have assessed IAT in patients who achieved successful recanalization following EVT, yet their findings have been inconsistent which suggests that IAT targeting microthrombi is insufficient for microcirculation reperfusion and requires multi-target intervention.

Adenosine confers a range of physiological benefits with mutiple target for AIS treatment. As a potent vasodilator, it enhances microvascular perfusion, suppresses neutrophil adhesion and migration, inhibits platelet aggregation, and reduces the generation of oxygen radicals^20^. Moreover, under low intracellular ATP conditions, adenosine exhibits cytoprotective effects by boosting ATP synthesis independently of its receptors, likely mitigating observed edema in endothelial cells and astrocyte end-feet^21^. Therapeutically, adenosine ameliorates cerebral ischemia-reperfusion injury by promoting microcirculatory reflow through pericyte relaxation and reducing ischemia-induced erythrocyte nesting, while potentially diminishing edema in endothelial cells and astrocytes to improve no-reflow^22^. Given the shared pathophysiology of no-reflow in cardiac and cerebral tissues, adenosine—already approved for reducing microvascular obstruction post-percutaneous coronary intervention (PCI)—shows promise for mitigating this phenomenon in stroke patients^23^. Therefore, this study will provide insight to explore the safety and efficacy of adjunctive intra-arterial adenosine administration in LVO stroke patients after successful EVT.

The REACT trial enrolled its first patient on November 13, 2024. The enrollment was finished on May 25, 2025. Full study completion (including collection on 3-month outcomes) is expected by August 2025. When completed, this trial will provide pivotal data allowing assessment of the safety and efficacy of intra-arterial adenosine in patients with acute ischemic stroke due to LVO following successful EVT.

## Data Availability

All data produced in the present study are available upon reasonable request to the authors

## Contributorship

ZZ, WZ, XH, DW, JLS, TNN, and RGN designed and conceptualized the study. XH, GL, XC and YH participated in data collection. XH, GL and XC wrote the manuscript. All authors critically revised and approved the manuscript.

## Acknowledgement

None.

